# Enhancing Uploads of Health Data in the Electronic Health Record – The Role of Framing and Length of Privacy Information: A Survey Study in Germany

**DOI:** 10.1101/2024.10.08.24315097

**Authors:** Niklas von Kalckreuth, Dr. Markus A. Feufel

## Abstract

**Background:** The German electronic health record (EHR) aims to enhance patient care and reduce costs, but users often worry about data security. In this article, we propose and test communication strategies to mitigate privacy concerns and increase EHR uploads.

**Objective:** We explore whether presenting a privacy fact sheet (PFS) before users must decide whether to upload medical reports in the EHR increases their willingness to do so. Our study examines the effects of framing and length of PFS on this decision.

**Methods:** In an online user study with 227 German participants, we used a realistic EHR click dummy and varied the PFS in terms of length (short vs. long) and framing (EHR-centered vs. patient-centered).

**Results:** The results show that a PFS has a positive effect on uploading (OR 4.276, P=.015). Although there was no effect regarding the length of a PFS, a patient-centered framing increased uploads compared to an EHR-centered framing (OR 4.043, *P*=.003).

**Conclusion:** Displaying PFSs at the beginning of an upload process is a cost-effective intervention to boost EHR adoption and increase uploads of medical reports. While the length of a PFS did not influence behavior, PFSs are maximally effective if they frame information in a way that emphasizes how users can exert control over their data. Willingness to upload medical data is key to the success of the EHR, including better treatments and lower costs.

## Introduction

### Background

The digital transformation of healthcare systems holds enormous potential for improving patient care while at the same time reducing costs.^1,2^ In this process, the electronic health record (EHR) – which was introduced in Germany in 2021 and was implemented nationwide and mandatory for all patients starting on January 15, 2025 unless they actively opt out – plays a key role.^3^ With the EHR patients’ health data (e.g. diagnoses, therapies, vaccinations, discharge reports, emergency data and medication plans) can be documented, exchanged and viewed.^4,5^ There are several benefits of the EHR on an individual level: For example, duplicate diagnoses could potentially be eliminated, and pre-existing conditions, intolerances and medication plans could be considered during treatment without the patient having to bring a stack of medical reports to the physician.^5^ In addition, using the EHR, physicians should have more time for the actual treatment of a patient, as they would spend less time on obtaining patient’s medical history.^6^ However, the success of the EHR depends on whether and under what circumstances patients will use it. In Germany and according to the Patient Data Protection Act, it is the patient alone who controls what data are stored, shared and displayed in the HER.^5^ Whereas 3 out of 4 Germans state in recent surveys that they would use the EHR^7^, all respondents in a recent interview study stated that they were concerned about the data security and data privacy of the EHR^6^, even though the German EHR adheres to the highest possible data security and privacy standards, which are higher than for most online banking applications.^8^ To ensure successful implementation of the EHR, users should thus be transparently informed about its privacy and data security standards. Ultimately, we suggest that knowledge of these standards may reduce privacy concerns and increase the likelihood of use.

### Related Work

‘Notice and choice’ is the most widely used model for ensuring data privacy worldwide.^9,10^ It comprises two main strategies: privacy notices and privacy choices. *Privacy notices* describe how personal data is collected, processed, and shared with third parties, whereas *privacy choices* give users control over various aspects of these practices, including the decision to start and terminate them.^9^ Various studies show that privacy notices help users make effective privacy choices, that is, identify and use systems with high privacy and data security standards and reject those that are less secure.^11,12^ The most frequently used privacy notices are *privacy policies*, which are public documents where companies describe their data collection and data use practices in detail.^13^ Since the introduction of the General Data Protection Regulation (GDPR) in 2018, privacy policies are mandatory for all providers that collect personal data in the EU.^14^ However, privacy policies are often not read by users, e.g., because they are long and use legal jargon, which is difficult to understand if not incomprehensible for most users.^15–19^ Thus, an alternative strategy to communicate data security and privacy standards is warranted if their goal is to inform patients transparently about the security of their data. Ultimately, more transparent privacy notices have been shown to increase user control^20^, trust in the provider^21,22^ and reduce privacy concerns and increase EHR use^23–25^. Research concerning with social network sites (SNS) indicates that such information may increase the perception of control, which in turn can lead to greater willingness to disclose personal and sensitive information.^26,27^

To create more user-friendly privacy notices, previous research suggested a simple list of questions that describe exactly what happens to the data.^28^ In contrast to privacy policies, such simple lists or *transparency features* use easy-to-understand language to provide a brief overview of relevant privacy practices.^29^ Whereas a study in the eCommerce domain demonstrated that transparency features positively influence purchase numbers^30^, another study with SNS showed more multifaceted results. Users’ decision to disclose their data depended on the privacy policies mentioned in the transparency feature, for instance, how well the data were protected and when the information was made available to users (e.g., the temporal proximity to the decision to disclose data).^31^

Although a transparency feature is meant to summarize essential information of privacy policies, it is unclear which information users consider essential. Some evidence to answer this question can be found in the *transparency for trust principle*, which summarizes three information needs of mHealth app users^32^ based on experimental studies, systematic reviews, and reports of patient concerns: a) what data are collected? b) how they are stored (e.g., anonymized or encrypted)? and c) who has access to the data?

In addition, there is some research as to how this information should be best communicated. First, it is stated that transparency features should be easy to understand and read.^33^ Second, both text length and word choice have been said to influence users’ data disclosure.^15,34,35^ Third, in our own study^36^, we created a privacy fact sheet (PFS) based on these recommendations and showed that a longer, more detailed text can increase users’ decision to upload their data to the EHR compared to a shorter, less detailed version, if provided directly before the upload decision. Fourth, the framing of information – the way information is presented to users^37^ – influences user’s decisions to disclose their data.^26,38,39^ For instance, a more specific framing of information categories (e.g., “Privacy Settings” versus “App Settings”)^26^ and a positive framing (e.g., “tag me” versus “do not tag me”)^39^ increased users’ willingness to disclose personal information in the context of SNS. Additionally, a social framing emphasizing peer behavior (e.g., “a majority of users accept cookies”) increased cookie acceptance.^38^

In the present article, we build on and extend previous findings by framing transparency features around the concept of data autonomy. Specifically, research has shown that users want control over their own (health)data^20,21,40^ and that perceived control may increase technology use.^23–25,31^ Thus, we investigate whether framing privacy information around how EHR users can control their data in and with the EHR increases their decisions to upload medical reports.

### Aim of this Research and Approach

In this study, we pursue two objectives: (1) we aim to replicate our previous finding that displaying a PFS at the point of data disclosure and longer rather than shorter texts make EHR uploads more likely, and we aim to extend these findings by examining whether (2) the framing of a PFS around what the EHR does (an EHR-centered framing) or what it allows its users to do (a patient-centered framing) has an additional effect on the likelihood of EHR use. To achieve these goals, we empirically investigate the influence of a long and a short PFS with either system-centered or patient-centered framing on EHR upload behavior. After discussing the results, we derive practical implications, reflect limitations of the study and provide a conclusion.

## Methods

### Ethics Approval and Consent to Participate

This study was approved by the Ethics Committee of the Department of Psychology and Ergonomics at Technische Universität Berlin (tracking number: AWB_KAL_02_230510_Erweiterungsantrag). Participants volunteered to participate in the survey, and written informed consent was required. On the first page of the survey, participants were told about the investigator, the study purpose, what data were to be collected during the study, and where and for how long they would be stored. Participants had the possibility to download a pdf providing the above information. Before they were asked for written informed consent, participants were informed about the duration of the survey (approximately 7 minutes) as well as the compensation for participation.

### Participants

The study was conducted between September 2 and September 16, 2023. The study was conducted between September 2 and September 16, 2023. To determine the necessary sample size, we performed an a priori power analysis for a logistic regression using G*Power (version 3.1.9.7), with the predictors text length (short vs. long) and framing (patient-centered vs. EHR-centered), a significance level of α = 0.05, a statistical power of 1–β = 0.80, and a conservative assumption of a 50% upload rate under the null hypothesis. Based on a pre-study with 80 participants, we further assumed the smallest expected effect size (of text length) to be an odds ratio (OR) of 2.32, which required a minimum sample size of n = 230 to be detected. We oversampled participants by 20-30% based on dropout rates observed in the pre-study, resulting in a planned total of n=295 participants.^36^ As the content and questions of the study were designed to fit the context of the German EHR, individuals aged 18 years and older residing in Germany were allowed to participate in the study. Sampling was conducted through Prolific, a click worker platform characterized by high data quality.^41^ Participation was compensated with 1.40€ (US $1.48) according to minimum wage. The mean participation time was 8:12 minutes (SD 3:22 minutes) and the median was 7:15 minutes.

### Design

We used a 2-factorial between-subjects design with the independent variables (IVs) text length (long vs. short) and text framing (patient-centered vs. EHR-centered) along with a control group (no PFS displayed). The five experimental conditions included (1) no PFS, (2) a short, EHR-centered PFS, (3) a long, EHR-centered PFS, (4) a short, patient-centered PFS and 5) a long, patient-centered PFS that were administered immediately before participants were asked to decide whether to upload a medical report to an EHR click dummy. Participants in the control group (no PFS) received no additional privacy information, reflecting the current practice of most German EHR applications. Participants were randomly assigned to one of these conditions in parallel (single-blinded, simple randomization, ratio: 1:1:1:1:1) using LimeSurvey’s built-in “rand” function. The dependent variable was the decision to upload the diagnosis, that is, whether participants were willing to upload the medical findings to the EHR.^42,43^

### Materials

Following a standard methods in technology acceptance studies, we used a case vignette to evoke a typical situation in which an EHR would be used.^44^ The vignette describes a scenario in which participants are first asked to imagine that they are suffering from moderate to severe depression and then to decide whether to upload this report to their EHR. We chose a stigmatized disease because uploading reports on such diagnoses is perceived as riskier, highlighting privacy issues^42,43,45^ and thus increasing the likelihood that participants will pay attention to privacy and data security notices. The case vignette can be found in the Appendix.

To create the PFSs, we identified content based on the “privacy and data security” category of the transparency for trust principles.^32^ According to these principles, privacy notices must specify which data leave the system, how they are stored and protected and who has access to the data. On this basis, we identified four categories of information related to data security, data control, storage duration and storage location. Content for each of these categories was created based on actual information on the German EHR. We formulated all texts following the guidelines for improving instructional and informational texts^35,46^, including unequivocal headings, standard rather than technical language and short summaries of the content of each category. In the end, each of the four information categories consisted of a category heading, a short summary and more detailed information. For the short version of the PFS, only the headings and the short summaries were displayed, whereas in the long version the more detailed information was displayed in addition to the headings and the summaries. Figure 1 shows the English versions of the PFSs used, comparing the short and long text formats.

**Figure 1.**
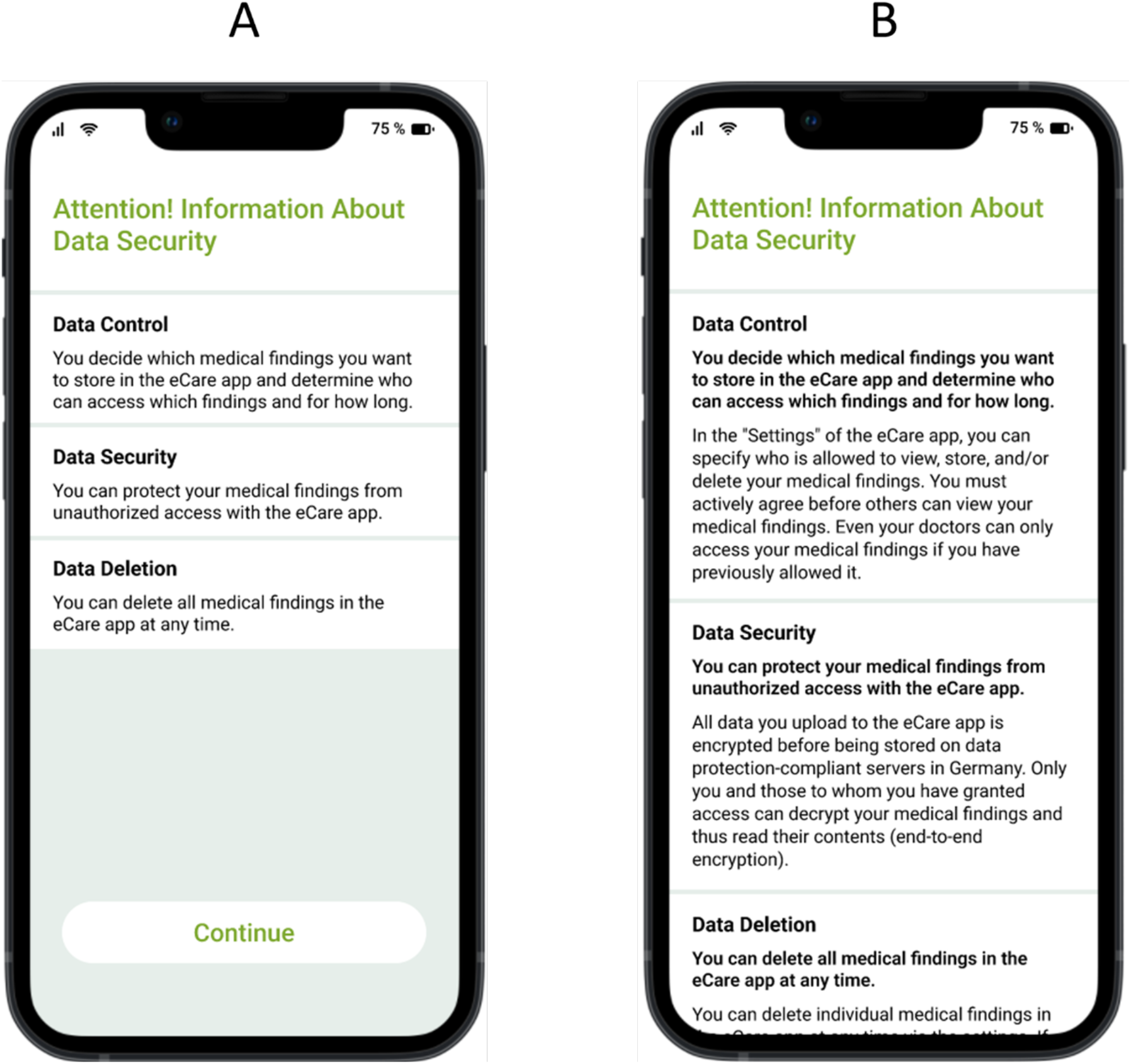
Privacy fact sheets, used in the study with (A) short and (B) long text formats.

To create a patient-centered framing of the PFS, the information was formulated in such a way that the users of the EHR app were actively addressed as actors and had a clear understanding of the extent to which they themselves can use the EHR to exercise control over their data (“You can protect your medical findings from unauthorized access with the eCare app”). For the variant of the PFS with an EHR-centered frame, the information was formulated in such a way that the EHR app is described as the main actor and control instance (“The eCare app protects your medical findings from unauthorized access”). Figure 2 shows the English versions of the PFSs, comparing the EHR-centered and patient-centered framing. The English translation of the full texts can be found in the Appendix.

**Figure 2.**
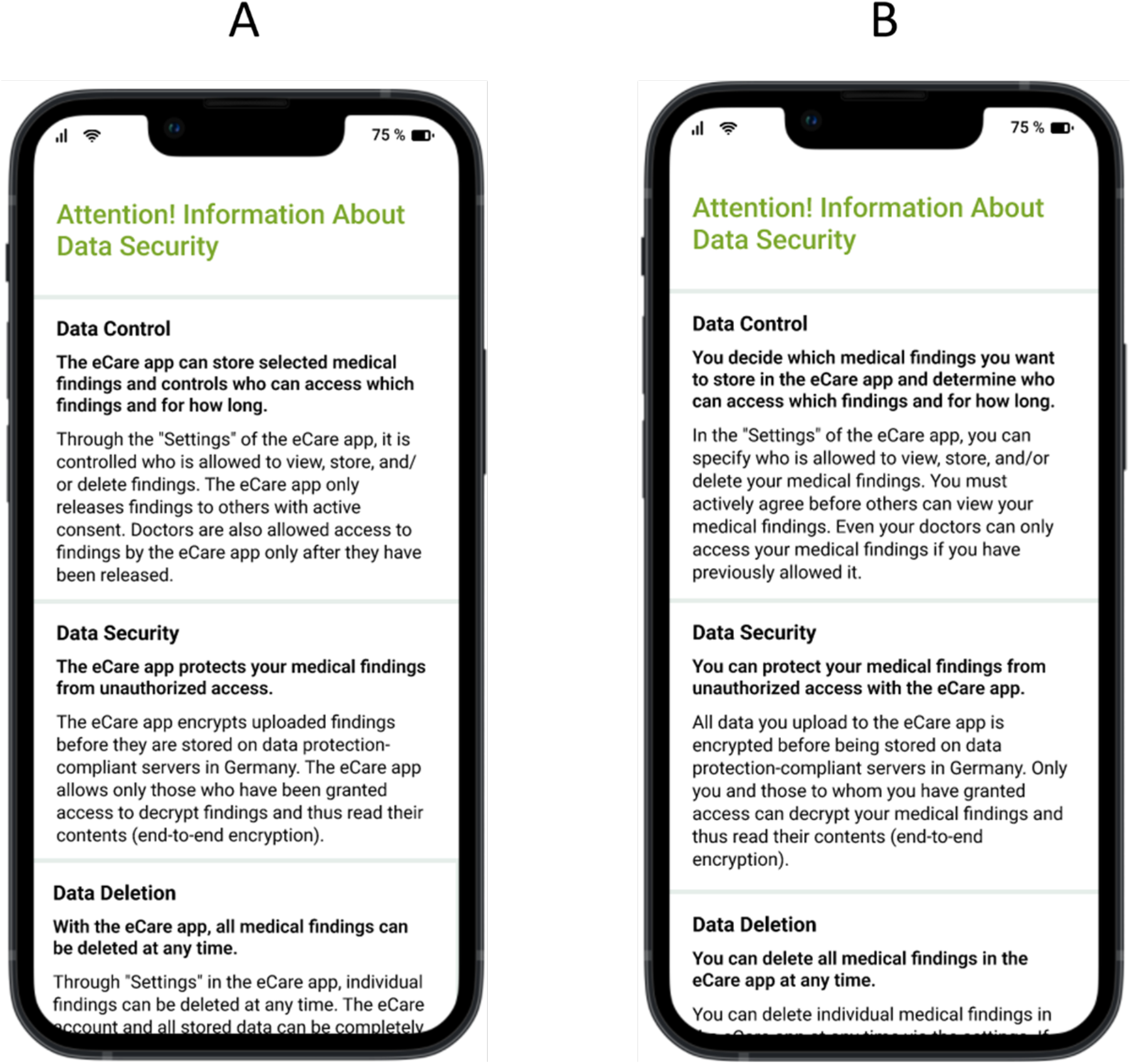
Privacy fact sheets, used in the study with (A) EHR-centered and (B) patient-centered framing.

For the study, an interactive functional prototype (a click dummy) was created using FIGMA (a software for interface design) and modeled after the mobile EHR application (the eCare app) of a German health insurance company (the BARMER). This prototype allows a realistic interaction with an EHR. Specifically, the prototype gave participants the ability to upload medical reports, to grant, or revoke permissions to view them, and to create medication plans. Only the “Upload findings” function was used in this study.

We used LimeSurvey (version 3.28.3+220315) to create and conduct an 8-page online survey. The EHR prototype was embedded in the survey using iFrame. LimeSurvey software was used to ensure that all questions had to be answered to complete the study and receive compensation. As manipulation checks, we queried perceived reading speed and perceived control over their data for each PFS, assuming a high perceived reading speed for a short length and a high perceived control over data for the text with patient-centered framing. The items for perceived reading speed and perceived control over data were self-constructed and pre-tested, following standard guidelines for the construction of items, such as the use of simple language, short, specific, and neutral wording (e.g., no leading questions or double negation) and one-dimensionality (i.e., each question refers to a single fact).^47,48^ The item on perceived reading speed has been validated in a previous study^36^, whereas the item on perceived control was newly developed for this study and pre-tested in a pilot study with 80 participants to ensure clarity and content validity. Both manipulation checks were measured using a 7-point Likert scale ranging from 1 (“Strongly disagree”) to 7 (“Strongly agree”). The decision to upload the finding was measured using a dichotomous item (yes/no), which has been validated in previous studies on EHR usage behavior.^36,42^ The full questionnaire can be found in the Appendix.

### Procedure

The study procedure is shown in Figure 3. The survey consisted of 4 parts. After giving their informed consent according to the WMA Declaration of Helsinki, participants read the case vignette (1). Then participants could interact with the click dummy of the EHR app (2). First, participants were asked to select a medical report based on the findings discussed in the case vignette as preparation for the uploading procedure (2a) and were randomly assigned to one of the five experimental groups (i.e., no PFS, short, EHR-centered PFS, long, EHR-centered PFS, short, patient-centered PFS and long, patient-centered PFS). As part of the uploading process, participants in the four intervention groups were then each shown the corresponding PFS (2b). Participants then decided whether they wanted to upload the report to their EHR (2c). Afterwards, participants in the experimental groups were asked questions about the content of the PSF to ensure that the text had been read (attention checks) and about the perceived reading speed and the perceived control over data (manipulation checks) (2d). Next, the questionnaire was answered by the participants (3). Finally, demographic characteristics (age, gender, education level, and experience with mHealth applications) were collected as control variables, and participants were given the opportunity to declare their responses invalid in case they did not pay sufficient attention to the instructions provided, while still receiving compensation (4).

**Figure 3.**
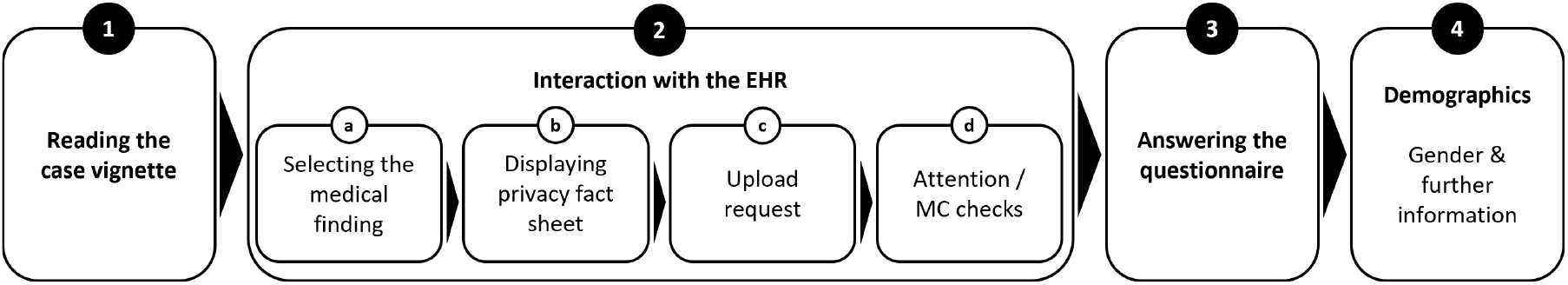
Overview of the study procedure.

### Hypotheses

In summary, we hypothesize that a short PFS will result in high perceived reading speed and the patient-centered framing in a high perceived control over data. We further hypothesize that the upload decision is positively influenced by showing a PFS versus no PFS (H1). More specifically, a long PFS will result in more uploads than a short PFS (H2) and a patient-centered framing will result in more uploads than an EHR-centered framing (H3).

### Analyses

We cleaned and analyzed the data using RStudio (version 1.3.1093). The influence of a PFS on the upload decision was tested using a generalized linear model (logistic regression with dummy coding). To test the hypotheses, we used planned contrasts. We adjusted p-values to mitigate multiple testing problems using the Benjamini-Hochberg procedure.^49^ The analysis regarding perceived reading speed and perceived control over data were performed using t-tests.

We also included a robustness check of the results: To control for potential influences of demographic and interindividual variables that could bias coefficients and p-values, we used multiple logistic regression.^50^ To not bias p-values as a result of controlling, we only included variables in the model that have been shown to have a causal relationship with the independent variable (i.e., causal confounders): age, education level, and experience with the mHealth systems.^21,51,52^ P-values were adjusted for multiple testing again using the Benjamini-Hochberg procedure.^49^

## Results

### Survey Characteristics

A total of 295 observations were collected, of which 68 records were excluded. Specifically, 20 records were excluded because of incomplete questionnaires, 45 because they failed attention checks, and three because they were marked as invalid by participants. Figure 4 shows how the trial was conducted and participants allocated to the intervention groups, and their records analyzed as a flow chart.

**Figure 4.**
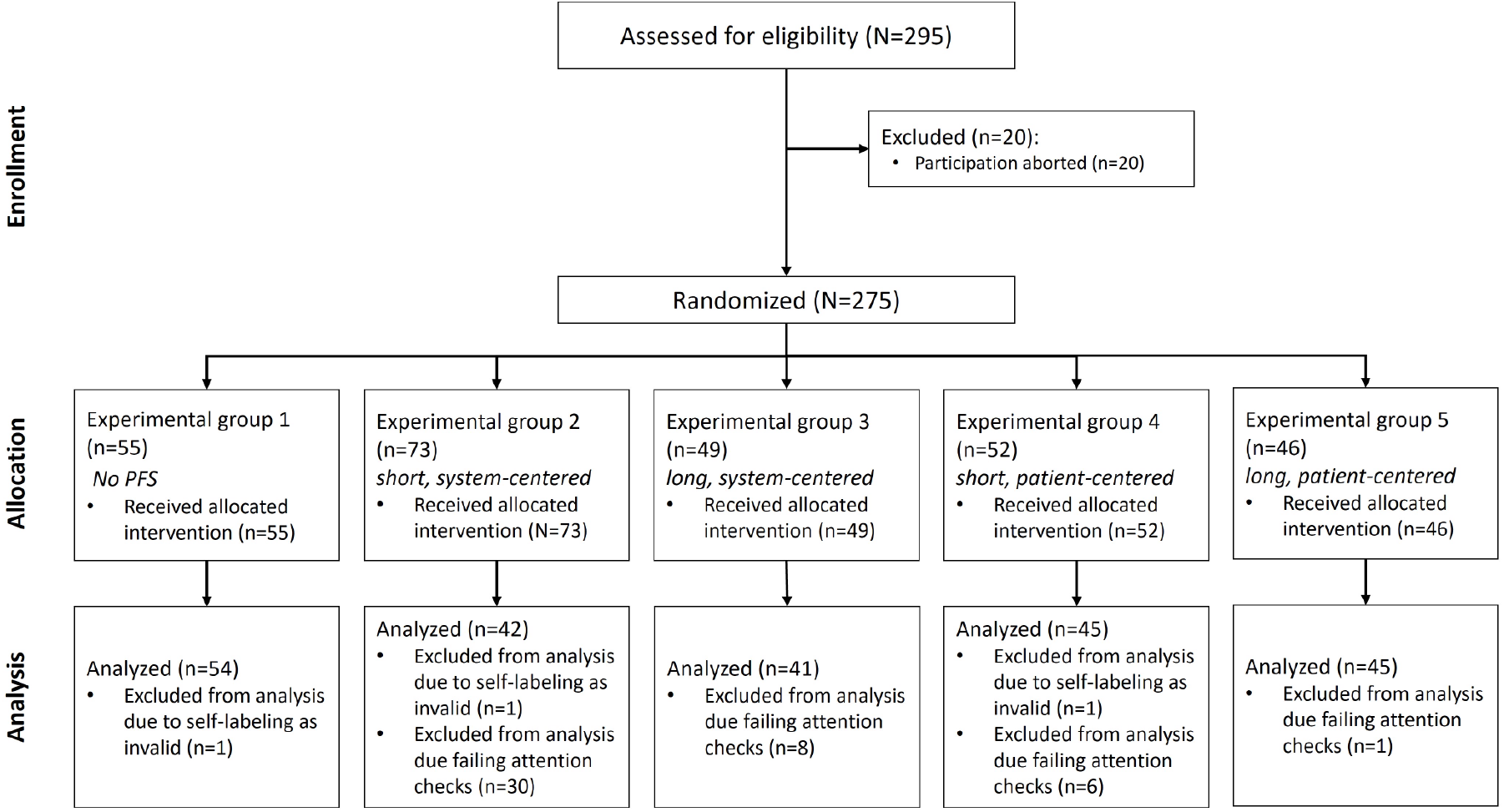
Flow chart of participant assignment to the experimental groups and exclusions.

The remaining sample of 227 observations (96 female, 125 male, 4 diverse, 2 no information) was used for further analysis. Table 1 summarizes the demographic characteristics of the sample.

**Table 1.** Demographic data of the sample (N=227).

| Demographic characteristics | No PFS<br>(n=54) | Short,<br>EHR-Cent<br>(n=42) | Long,<br>EHR-Cent<br>(n=41) | Short,<br>Pat-Cent<br>(n=45) | Long, Pat-Cent<br>(n=45) | Total<br>(N=227) |
| --- | --- | --- | --- | --- | --- | --- |
| Age (years), mean (SD) | 29.91<br>(8.84) | 29.9<br>(8.89) | 32.27<br>(9.35) | 29.56<br>(10.16) | 29.18<br>(7.5) | 30.12<br>(9.04) |
| **Gender, n (%)** |  |  |  |  |  |  |
| female | 27 (50) | 21 (50) | 14 (34.1) | 16 (35.6) | 18 (40) | 96 (42.3) |
| male | 26 (48.1) | 20 (47.6) | 26 (63.4) | 27 (60) | 26 (57.8) | 125 (55.1) |
| diverse | 1 (1.9) | 1 (2.4) | 0 (0) | 1 (2.2) | 1 (2.2) | 4 (1.8) |
| no answer | 0 (0) | 0 (0) | 1 (2.4) | 1 (2.2) | 0 (0) | 2 (0.9) |
| **Education, n (%)** |  |  |  |  |  |  |
| Highschool / vocational education | 24 (44.4) | 22 (52.4) | 17 (41.5) | 26 (57.8) | 19 (42.2) | 108 (47.6) |
| Bachelor degree | 16 (29.6) | 10 (23.8) | 9 (22) | 13 (28.9) | 13 (28.9) | 61 (26.9) |
| Graduate degree or higher | 14 (25.9) | 10 (23.8) | 15 (36.6) | 6 (13.3) | 13 (28.9) | 58 (25.6) |
| **Experience with mHealth apps, n (%)** |  |  |  |  |  |  |
| No use | 28 (51.9) | 19 (45.2) | 21 (51.2) | 27 (60) | 25 (55.6) | 120 (52.9) |
| Regular use | 26 (48.1) | 23 (54.8) | 20 (48.8) | 18 (40) | 20 (44.4) | 107 (47.1) |

Note: EHR-Cent=EHR-centered, Pat-Cent=patient-centered.

### Perceived Reading Speed and Perceived Control over Data

The perceived reading speed and perceived control over data served as a manipulation check to test our manipulations (see Figure 5). As expected, the reading speed was perceived as significantly faster when given the short PFS (M 6.41, SD 1.08) than the PFS with long text format (M 4.64, SD 1.31; *t171*= 9.71, *p*<.001). In addition, the perceived control over data was significantly higher for the PFS with patient-centered framing (M 6.5, SD 0.9) than EHR-centered framing (M 4.66, SD 1.21; *t171*= −11.363, *p*<.001). Consequently, we assume that our manipulations were successful.

**Figure 5.**
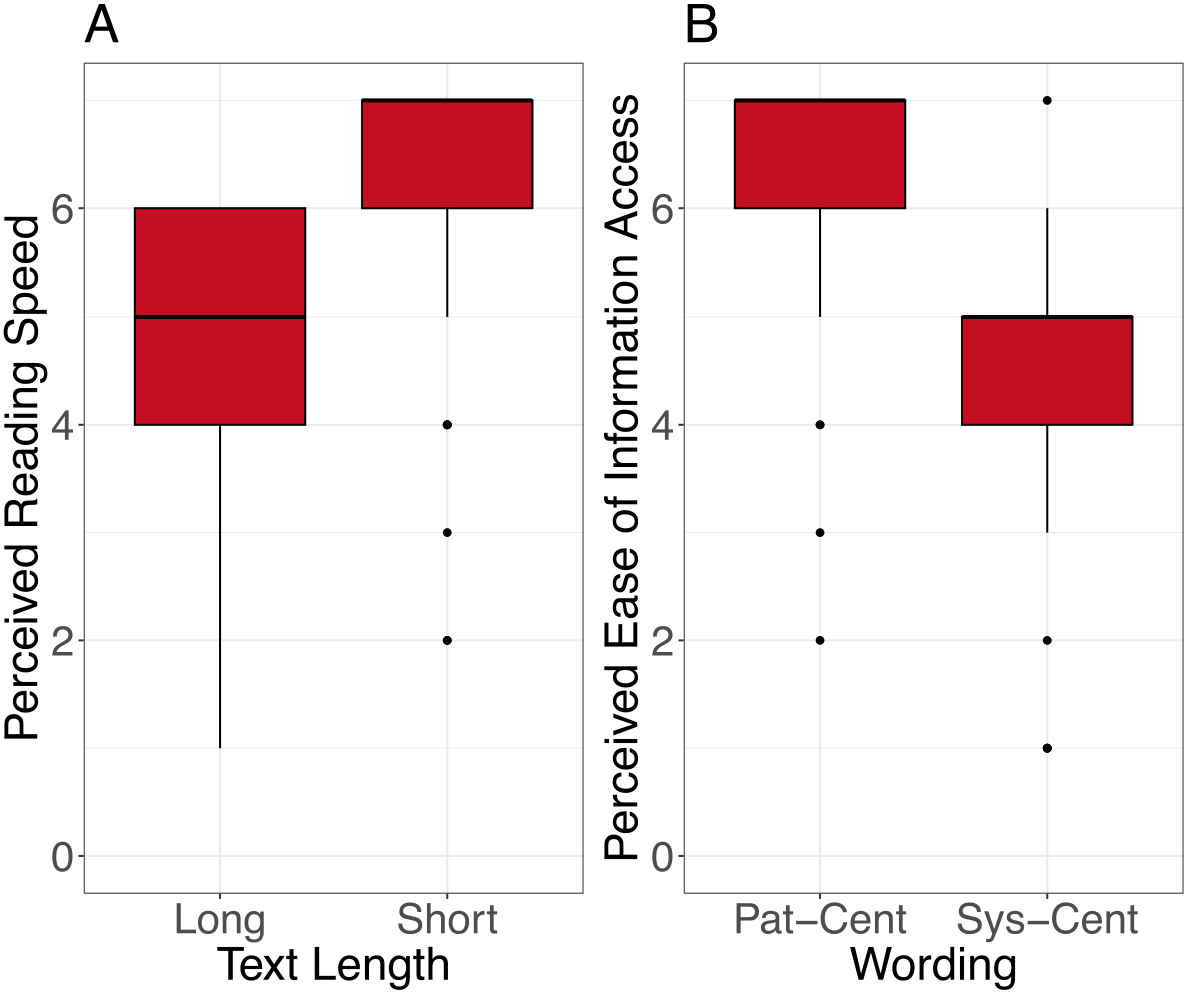
(A) Perceived reading speed concerning the length (short, long) and (B) perceived data security control concerning the framing (EHR-centered, patient-centered) of the PFS. The horizontal line in the box represents the median. Note: Pat-Cent = patient-centered and EHR-Cent = EHR-centered.

### Upload Behavior

Displaying a PFS generally increased uploading behavior (*z*=2.444, *p*=.015), thus supporting H1. Specifically, when participants received the PFS, they were four times as likely to upload the report to the EHR (OR 4.276; 95% CI 1.333-13.717). There was no significant difference in the upload behavior regarding the length of the PFS (*z*=1.821, *p*=.069). Consequently, H2 is rejected. A PFS with patient-centered framing increased upload behavior compared to the EHR-centered framing (*z*=2.928, *p*=.003), thus supporting H3. Participants seeing the PFS with patient-centered framing were four times more likely to upload the report to the EHR compared to participants seeing a PFS with EHR-centered framing (OR 4.043; 95% CI 1.587-10.301). The summary of the results of the logistic regression are shown in Table 2. The proportion of uploads by PFS framing and text length is shown in Figure 6.

**Table 2.**
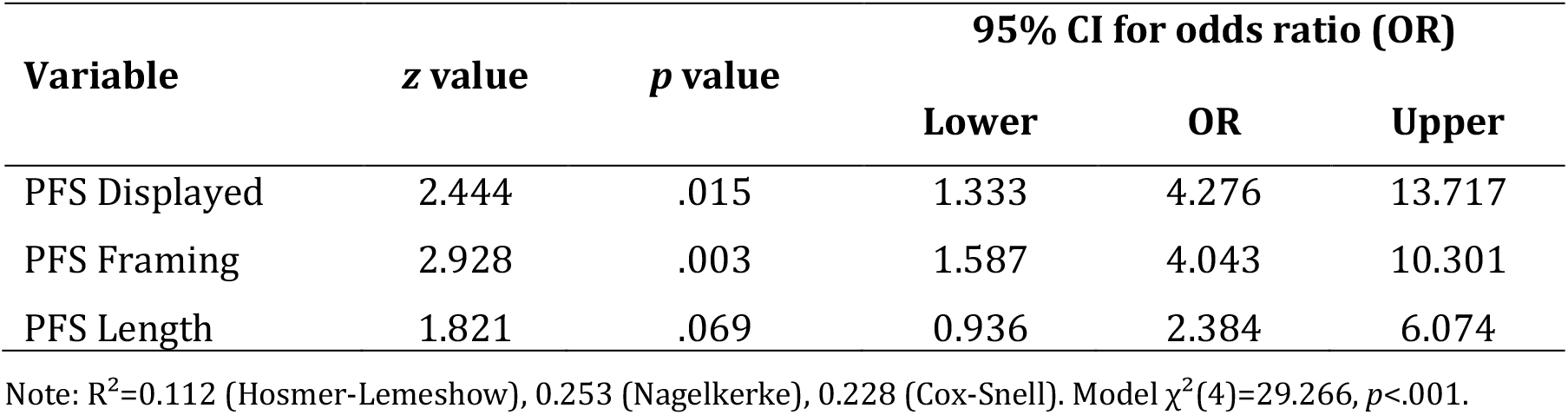
Results of the logistic regression.

| Variable | z value | p value | 95% CI for odds ratio (OR) |  |  |
| --- | --- | --- | --- | --- | --- |
|  |  |  | Lower | OR | Upper |
| PFS Displayed | 2.444 | .015 | 1.333 | 4.276 | 13.717 |
| PFS Framing | 2.928 | .003 | 1.587 | 4.043 | 10.301 |
| PFS Length | 1.821 | .069 | 0.936 | 2.384 | 6.074 |

Note: $R^2=0.112$ (Hosmer-Lemeshow), 0.253 (Nagelkerke), 0.228 (Cox-Snell). Model $\chi^2(4)=29.266, p<.001$ .

**Figure 6.**
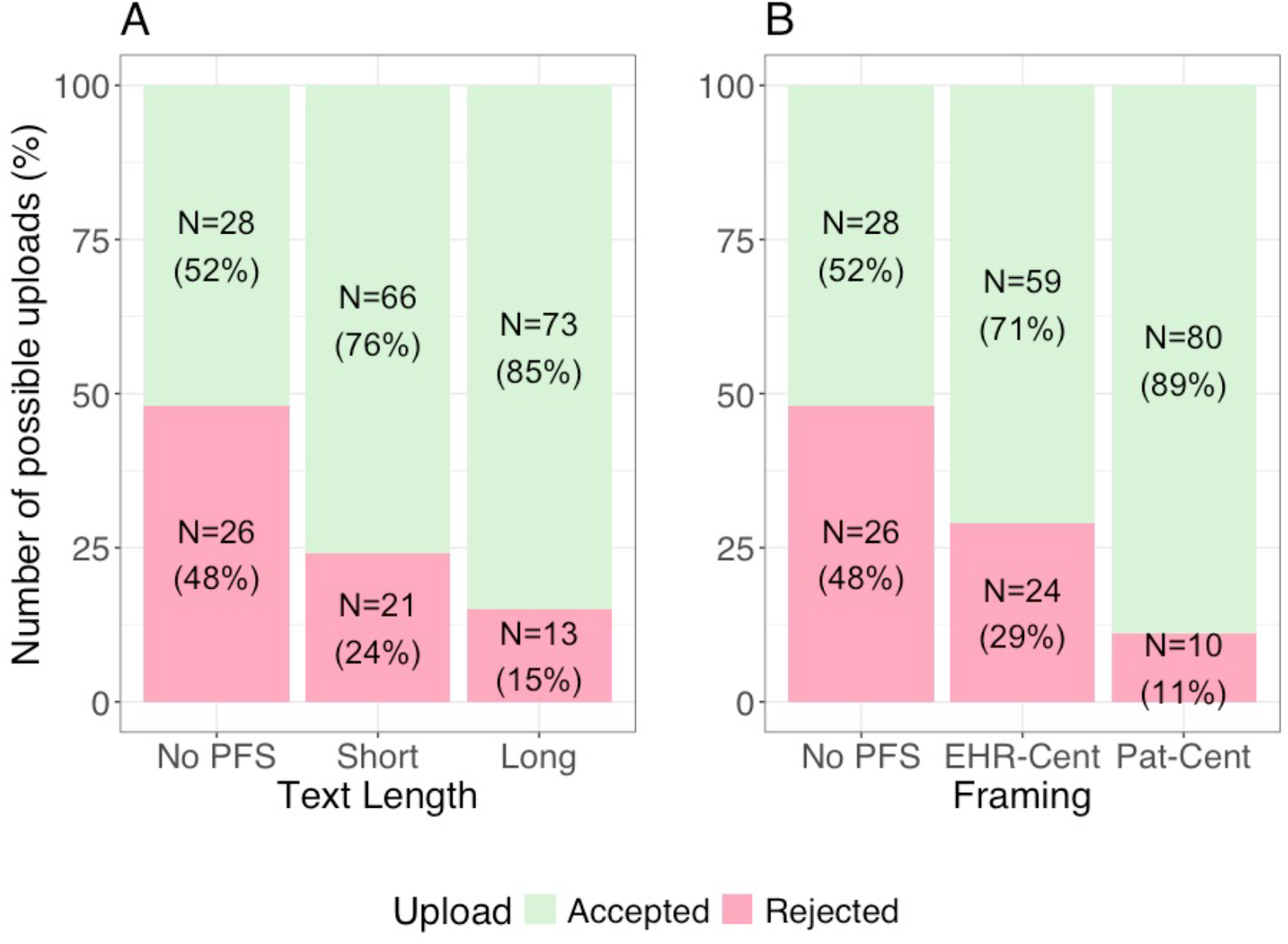
Number of uploads to the EHR in relation to the (A) text length (short, long) and (B) framing (EHR-centered, patient-centered) of a PFS. Note: EHR-Cent=EHR-centered, Pat-Cent=patient-centered.

### Robustness Check

When controlling for interindividual variables, the results remained stable. Showing a PFS still increased uploading behavior after controlling for all causal confounding variables (*z*=2.435, *p*=.023) as did the patient-centered compared to the EHR-centered framing (*z*=2.802, *p*=.005). In this model, gender, age, education level, and prior EHR use were included as covariates. Participants who selected ‘diverse’ (n = 4) or ‘no answer’ (n = 2) for gender were excluded due to small cell sizes. In summary, the control variables did not influence upload behavior.

## Discussion

### Principal Findings

The results of our study show that displaying a PFS has a positive influence on EHR use, specifically, the likelihood that users upload sensitive health data. When users saw a PFS shortly before they were asked to upload a stigmatized diagnosis to their EHR, they were 4 times as likely to upload this medical finding. This finding contrasts with previous studies on the effect of PFSs, which showed little, no, or even a negative impact of PFS on user behavior in non-medical domains, such as online shops, social networks, and booking portals.^29,31^ This difference may suggest that, to be effective, privacy notes must be framed around privacy practices that align with users’ interests in the domain at hand.^53^ For instance, the domains considered in previous studies emphasize economic interests in user data. Even if related practices are transparently communicated, users may (still) disagree with them.^29^ In contrast, in our study the PFS emphasized patient-centered use and control over health data that are to be used for one’s own benefit. Unlike previous studies that tested positive or negative affective framing – with mixed results regarding their impact on disclosure behavior and privacy awareness^31,54,55^ – our findings suggest that emphasizing how users may exert control over their data through a patient-centered framing may be a more effective strategy to support informed privacy decisions.

A surprising finding is that the framing but not the length of the PFS influenced upload behavior, which contrasts with our previous findings.^36^ When PFS framing and length are compared, what seems to count for the decision to upload health data to the EHR is users’ perceived control over their data^20,21,23,31^, which was maximized using a patient-centered framing of privacy control policies (see Figure 5B). That is, users’ willingness to engage with the EHR increases if they information about what they can do to control their data privacy and security rather than lengthy descriptions of what the EHR app can do for them. Thus, our study demonstrates that even a short but well-calibrated framing can enhance patient’s perceived control over their health data, which in turn can significantly impact their willingness to upload medical findings into the EHR. This underscores the importance of carefully designing and testing the presentation of privacy information to align with the specific expectations and needs of users in the context of health data management.

### Implications

The opportunities offered by implementing PFSs in the EHR should be considered by healthcare stakeholders. To date, privacy information in German EHR systems is typically limited to FAQ pages or static privacy policies that are not integrated into the user interface at the point of disclosure. Our study shows that PFSs can not only reduce general privacy concerns but may also address disease-related concerns, for instance, related to stigma.^42,43^ A patient-centered compared to an EHR-centered framing is perceived as providing greater control over one’s data and increases users’ willingness to upload medical reports into the EHR. If implemented on a large scale, the illnesses, allergies and medications of more patients could be considered in diagnostics and therapies. In this context, privacy fact sheets could serve as a practical tool in both interface design and policy implementation to support informed decision-making and strengthen digital trust in privacy-sensitive technologies. This, in turn, could lead to more efficient processes and reduce costs in the healthcare system. Furthermore, extensive health data sets increase the likelihood that treating physicians can diagnose rare diseases by accessing the experiences and diagnoses of other clinicians.^2^

### Limitations and Future Directions

There are several limitations of our study, which need to be considered in subsequent studies. Previous survey studies report that the adoption and approval of data-gathering technologies are significantly influenced by cultural factors.^56^ In comparison to citizens of other European countries, the German population is said to be particularly cautious when disclosing personal information online^57^, yet 93% of the population are internet users.^58^ Given that data were collected solely from German residents, future studies should validate the applicability of these findings with studies in other countries to help better understand the role of cultural differences. Moreover, privacy fact sheets (PFS) could be adapted to reflect the legal frameworks, technical infrastructures, and privacy expectations in different countries and tested accordingly to evaluate their cross-cultural effectiveness. Hence, contextual factors such as levels of digital trust or the structure of national healthcare systems may influence how users respond to privacy communication in different countries.

Furthermore, in the case vignette, we used a stigmatized illness (depression) to make the interaction riskier and thus focus on privacy issues. However, this approach limits the generalizability of our findings. Future studies should test the applicability of PFSs across a range of diseases with varying disease-related stigma, as the degree of perceived stigma may moderate the effectiveness of privacy communication interventions. In order to further strengthen the validity and generalizability of our results, two follow-up studies should examine the perspectives of people who (1) are already affected and those who (2) are not affected and compare them with the results of this study.

Moreover, due to the small number of participants who identified as ‘diverse’ or chose not to disclose their gender, these individuals had to be excluded from the regression analysis. This limits the generalizability of our findings to gender-diverse populations and underscores the importance of more inclusive sampling in future research.

Additionally, our study faced limitations due to uncontrolled conditions like participant’s location and potential distractions, as participants completed the questionnaire online. Future research could validate our findings through a laboratory study, ensuring a more controlled environment.

Finally, although a realistic click dummy was used, ta survey study hardly allows participants to immerse themselves. In a follow-up field study, researchers could collaborate with health insurers to gather real-world data on uploading behavior with an actual EHR. Conversely, our online study could not control for the situation in which participant answered our questions. Future research should validate our findings also in a lab study, ensuring a more controlled environment.

## Conclusions

Our results show that displaying a PFS to EHR users may increase their decisions to upload medical reports to the EHR. Specifically, our findings indicate that the patient-centered framing, specifying how users can control their data with and in the EHR app, increases user perceptions of control over their data and boosts the likelihood of EHR uploads, whereas the length of the PFS did not. This suggests that the effectiveness of PFS in enhancing user engagement is more dependent on how the information is presented than on the quantity of information provided.^36^ The implementation of PFS in user interfaces for privacy-sensitive technologies may represent a practical response to user needs for control and transparency. In the context of EHRs, for example, displaying PFSs with a patient-centered framing as part of the upload process could be an inexpensive but effective intervention to increase the EHR uptake and technology acceptance – particularly among users with privacy concerns (e.g., as in Germany). Ultimately, this could help ensure that more patients enjoy the benefits of these systems (e.g., more efficient healthcare processes, improved treatment outcomes, and reduced costs) and thus promote health equity.

## Supporting information

Case vignette

Privacy Fact Sheets

Questionnaire

## Data Availability

All data produced in the present study are available upon reasonable request to the authors

## Abbreviations

EHR: Electronic Health Record
EHR-Cent: Electronic Health Record centered
GDPR: General Data Protection Regulation
IV: Independent variable
M: Mean
mHealth: Mobile health
OR: Odds ratio
Pat-Cent: Patient centered
PFS: Privacy fact sheet
SD: Standard deviation
SNS: Social network sites

## Acknowledgements

We acknowledge support from the German Research Foundation and the Open Access Publication Fund of TU Berlin. We also thank the evangelisches Studienwerk Villigst and the German Federal Ministry of Education and Research, who provided the doctoral scholarship (NvK) without which this research would not have been possible. We thank all those who participated in the study.

## Contributorship

NvK researched literature and conceived the study under the supervision of MAF. NvK was involved in protocol development, gaining ethical approval, patient recruitment, and data analysis. NvK wrote the first draft of the manuscript. All authors reviewed and edited the manuscript and approved the final version of the manuscript.

## Declaration of conflicting interests

The authors declared no potential conflicts of interest with respect to the research, authorship, and/or publication of this article.

## Funding

Not applicable

## Ethics approval

The Ethics Committee of the Department of Psychology and Ergonomics (Institut für Psychologie und Arbeitswissenschaft) at Technische Universität Berlin approved this study (tracking number: AWB_KAL_02_230510_Erweiterungsantrag). Participants volunteered to participate in the survey, and informed consent was required.

## Guarantor

NvK

## Notes

### Competing Interest Statement

The authors have declared no competing interest.

### Funding Statement

This study did not receive any funding

### Author Declarations

The Ethics Committee of the Department of Psychology and Ergonomics (Institut fuer Psychologie und Arbeitswissenschaft) at Technische Universitaet Berlin approved this study (tracking number: AWB_KAL_02_230510_Erweiterungsantrag).

### Summary of Updates

Adding a sentence to the abstract conclusion regarding the non-effect of the length on upload behavior. Adding more precisely implications in the implications and conclusion section

