## Supplementary material for "Enhancing Uploads of Health Data in the Electronic Health Record – The Role of Framing and Length of Privacy Information: A Survey Study in Germany": Case vignette

**Case Vignette (English translation)**

Imagine the following scenario:

1. you have recently started using your health insurer's electronic health record (EHR), the "eCare" app, to manage your medical data and records and share them with your physicians if you wish.

2. you have been suffering from moderate to severe depression* for several years.

3. you are now faced with the decision of whether or not to include the diagnosis of this illness in your EHR.

*Depression is a mental illness that can manifest itself in numerous symptoms. A persistently depressed mood, inhibition of drive and thinking, loss of interest and a variety of physical symptoms, ranging from insomnia to appetite disorders and pain, are possible signs of depression. The majority of those affected have suicidal thoughts sooner or later, and 10 to 15% of all patients with recurring severe depressive phases die by suicide. Once the correct diagnosis has been made, the situation is anything but hopeless. In recent decades, a lot has been done in terms of treatment and more than 80% of sufferers can be helped permanently and successfully.
