## Supplementary material for "Enhancing Uploads of Health Data in the Electronic Health Record – The Role of Framing and Length of Privacy Information: A Survey Study in Germany": Privacy Fact Sheets

### PFS1 – short, system-centered

#### Attention! Information About Data Security

**Data Control**

The eCare app can store selected medical findings and controls who can access which findings and for how long.

**Data Security**

The eCare app protects your medical findings from unauthorized access.

**Data Deletion**

With the eCare app, all medical findings can be deleted at any time.

### PFS2 – long, system-centered

#### Attention! Information About Data Security

##### Data Control

**The eCare app can store selected medical findings and controls who can access which findings and for how long.**

Through the "Settings" of the eCare app, it is controlled who is allowed to view, store, and/or delete findings. The eCare app only releases findings to others with active consent. Doctors are also allowed access to findings by the eCare app only after they have been released.

##### Data Security

**The eCare app protects your medical findings from unauthorized access.**

The eCare app encrypts uploaded findings before they are stored on data protection-compliant servers in Germany. The eCare app allows only those who have been granted access to decrypt findings and thus read their contents (end-to-end encryption).

##### Data Deletion

**With the eCare app, all medical findings can be deleted at any time.**

Through "Settings" in the eCare app, individual findings can be deleted at any time. The eCare account and all stored data can be completely deleted. To do this, consent to the privacy and terms of use must be revoked via email to.

### PFS3 – short, patient-centered

#### Attention! Information About Data Security

**Data Control**

You decide which medical findings you want to store in the eCare app and determine who can access which findings and for how long.

**Data Security**

You can protect your medical findings from unauthorized access with the eCare app.

**Data Deletion**

You can delete all medical findings in the eCare app at any time.

### PFS4 – long, patient-centered

#### Attention! Information About Data Security

##### Data Control

**You decide which medical findings you want to store in the eCare app and determine who can access which findings and for how long.**

In the "Settings" of the eCare app, you can specify who is allowed to view, store, and/or delete your medical findings. You must actively agree before others can view your medical findings. Even your doctors can only access your medical findings if you have previously allowed it.

##### Data Security

**You can protect your medical findings from unauthorized access with the eCare app.**

All data you upload to the eCare app is encrypted before being stored on data protection-compliant servers in Germany. Only you and those to whom you have granted access can decrypt your medical findings and thus read their contents (end-to-end encryption).

##### Data Deletion

**You can delete all medical findings in the eCare app at any time.**

You can delete individual medical findings in the eCare app at any time via the settings. If you no longer wish to use eCare at all, you can completely delete your eCare account and all your medical findings. Simply revoke your consent to the privacy and terms of use by email to.
