## Supplementary material for "Enhancing Uploads of Health Data in the Electronic Health Record – The Role of Framing and Length of Privacy Information: A Survey Study in Germany": Questionnaire

**Manipulation check (perceived reading speed):** Rate the following statement: the data security information can be read quickly. (self-constructed)

**Manipulation check (perceived data security control):** Please rate the following statement: The information on data protection made me aware that I, as a user, can effectively control the security of my data in the eCare app. (self-constructed)

**Attention check I:** Please answer the following question regarding the content of the privacy information displayed in the click dummy: Who has access to the records stored in the eCare app? (Query as multiple choice) (self-constructed)

**Attention check II**: Please answer the following question regarding the content of the privacy information displayed in the click dummy: At what point can the stored records be deleted? (Query as multiple choice) (self-constructed)

**Behavioral decision:** Would you like to upload the report to your electronic health record? (y/n) (von Kalckreuth et al., 2023)

**Demographics**

**Age**: Please enter your age.

**Gender (m/f/d):** Please indicate your gender.

**Education**: Please enter your highest qualification.

- No degree
- School leaving certificate
- Secondary school certificate
- General qualification for university entrance
- Vocational training
- University degree (bachelor's or master's)
- other

**Experience with mHealth apps**: How often do you use health or fitness apps?

- Never
- Tried once
- One app regularly
- Multiple apps regularly

**Reability Check:** Is there any reason why we should NOT use your data? You will be paid regardless of your response.

- Yes, I rushed through.
- Yes, I did not really read the questions.
- Yes, I choose random answers.
- Yes, for other reasons.
- No, you can use my data.
